# AI-Assisted Interpretation of Neurologic Autoantibody Cell-Based Assays

**DOI:** 10.64898/2026.10.16.26365578

**Authors:** Zhihui Qi, Zijian Cao, Hongyi Tan, Boxiang Tang, Samuel X Shi, Ruoyong Jia, Yang Yao, Jingshan Chen, Zongjiu Zhang, Jiahao Wan, Pei Zheng

**Affiliations:** Departments of Neurology, Tianjin Medical University General Hospital, Tianjin 300052, China; Departments of Neurology, Beijing Tiantan Hospital, Capital Medical University, Beijing, 100071, China; School of Biomedical Engineering, Tsinghua University, Beijing 100084, China; Jewelry and Metal Arts Program, California College of the Arts, 145 Hooper Street, San Francisco, CA 94107, USA; Alfred E. Mann Department of Biomedical Engineering, Viterbi School of Engineering, University of Southern California, Los Angeles, CA 90089, USA; Clinical Laboratory, Tianjin Medical University Cancer Institute and Hospital, Tianjin 300202, China; Institute for Hospital Management of Tsinghua University, Tsinghua University, Shenzhen 518052, China

## Abstract

Neurologic autoantibody cell-based assay images are difficult to interpret when fluorescence is weak, focal or dominated by background signal. We developed a visual artificial intelligence overlay that transfers detected cell boundaries from the structural channel to the antibody-staining image and evaluated standalone model concordance and its association with reader classifications. The analysis used 2 763 records for model development and 499 held-out records spanning 14 antibody targets. Four readers assessed the held-out records first without assistance and then with the boundary overlay and model category available. The selected model agreed with the reader-derived three-class reference in 85.2% of records, but recall was lowest for weak-positive or equivocal records. Reader agreement increased in the assisted session, and almost all revised ratings moved towards the model category. The combined display may support consistent visual review while also creating a risk of model-directed anchoring; its role as a second-read aid requires independent validation.

## Introduction

Autoantibody testing provides essential laboratory evidence for autoimmune neurological diseases, including autoimmune encephalitis, neuromyelitis optica spectrum disorder, myelin oligodendrocyte glycoprotein antibody-associated disease, autoimmune nodopathies, and paraneoplastic neurological syndromes^1,2^. By expressing target antigens in cultured cells and preserving their native conformational epitopes, cell-based assays (CBAs) are considered the reference method, and for many neuronal surface antibodies the diagnostic gold standard, in current clinical practice and diagnostic recommendations^3,4^. Nevertheless, although strong and characteristic fluorescence patterns are generally straightforward to recognize, weak, focal, or background-dominant staining can make borderline results highly dependent on reader judgment. CBA interpretation requires readers to determine whether fluorescence is specifically associated with antigen-expressing cells, follows the expected antigen-specific distribution, is sufficiently distinguishable from background and non-transfected cells, and is consistent across representative microscopic fields and assay controls. These judgments may be particularly difficult when only a small proportion of cells shows specific staining or when nonspecific fluorescence obscures the expected pattern. Consequently, neural autoantibody results may vary according to reader experience, specimen type, fixation method, substrate preparation, image-acquisition conditions, and detection platform^5,6,7,8,9^. Computer-assisted immunofluorescence may improve analytical standardization^10^, whereas multireader studies have demonstrated substantial experience-related variability in neural antibody interpretation^11^.

We developed a visual artificial intelligence (AI) overlay that maps cell boundaries detected in the green channel onto the corresponding red antibody-staining image. We evaluated two questions: how closely the selected classifier agreed with a reader-derived three-class reference, and how reader classifications changed when the overlay and model category were available during a second reading session. The study architecture and datasets are summarized in Fig. 1 and Table 1, and the technical pipeline is shown in Supplementary Fig. S1.

**Fig. 1.**
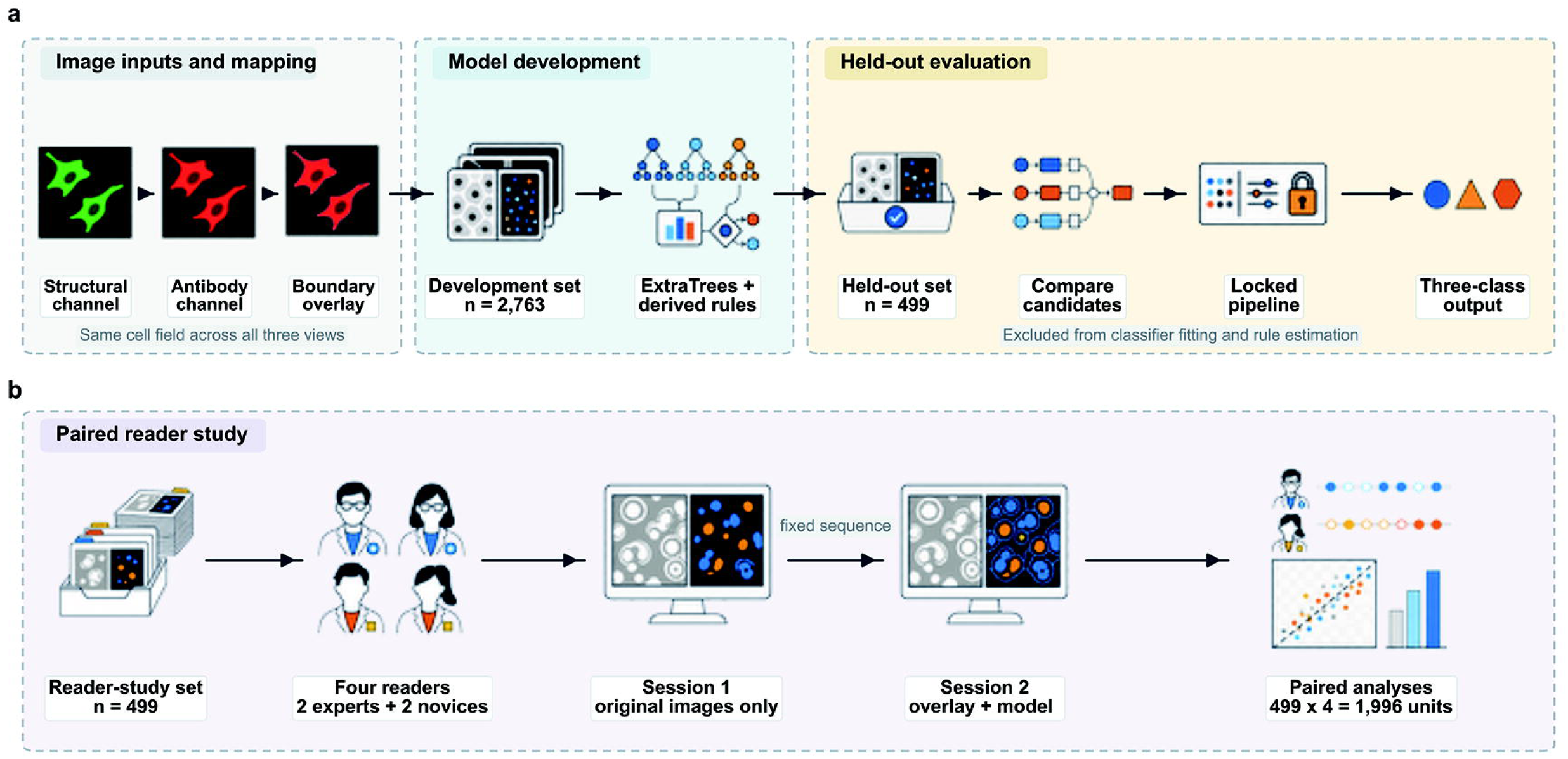
Study architecture. a, Paired cell-based assay (CBA) structural-and antibody-channel images were used to localize cells and transfer cell boundaries to the corresponding antibody image. The development set comprised 2 763 records. Candidate models were compared in a separate held-out set of 499 records before the final ExtraTrees-based pipeline and deterministic decision rules were locked. b, The paired reader study used the same 499 held-out records. Four readers (two experts and two novices) evaluated the original images in session 1 and repeated the assessment with the boundary overlay and model category available in session 2, yielding 1 996 paired reader-record observations. The performance reference was derived from session-1 ratings.

**Table 1.** Study datasets and analytical purpose. The held-out set was used for candidate-pipeline comparison, standalone model evaluation, and the paired reader study; it was not a strictly untouched external test set.

| Dataset | Records | Three-class composition | Analytical purpose |
| --- | --- | --- | --- |
| Model-development dataset | 2 763 | Negative, 1 509; weak positive/equivocal, 305; positive, 949 | Model fitting, cross-validation, and rule estimation |
| Held-out model-selection and reader dataset | 500 identified; 499 included | Negative, 258; weak positive/equivocal, 73; positive, 168 | Model selection/evaluation and two-session reader study |

## Materials and methods

### Study design and datasets

This retrospective analytical study comprised model development, held-out model selection/evaluation, and a two-session paired reader study. The unit of analysis was one paired-channel CBA record containing a green structural/cell-identification image and the corresponding red antibody-staining image. The development dataset contained 2 763 records. Of 500 records identified for held-out evaluation, one incomplete record was excluded, leaving 499 evaluable records (Fig. 1 and Table 1). Record selection aimed to balance category representation while considering the distribution encountered in clinical practice, prioritizing clear, evaluable paired-channel images.

The 499-record held-out set covered 14 antibody targets: acetylcholine receptor (AChR; n=45), adenylate kinase 5 (AK5; n=5), amphiphysin (n=15), aquaporin 4 (AQP4; n=45), contactin-1 (CNTN1; n=44), gamma-aminobutyric acid type B receptor (GABABR; n=45), glutamic acid decarboxylase 65 (GAD65; n=45), glial fibrillary acidic protein (GFAP; n=45), Hu antigen (n=15), leucine-rich glioma-inactivated protein 1 (LGI1; n=45), myelin oligodendrocyte glycoprotein (MOG; n=45), N-methyl-D-aspartate receptor (NMDAR; n=45), SRY-box transcription factor 1 (SOX1; n=45), and titin (n=15).

Clinical specimens and corresponding CBA images were provided by the neuroimmunology laboratories of Tianjin Medical University General Hospital and Beijing Tiantan Hospital, Capital Medical University. The classifier, thresholds, and post-processing rules were fitted using the development dataset; the 499-record held-out set was then used to compare candidate pipelines and select the final analysis model.

### Image acquisition and processing

Each record comprised paired green-and red-channel fluorescence images acquired during routine CBA testing. The green channel represented antigen-transfected cells carrying a green fluorescent protein (GFP) marker and was used to identify the corresponding cellular structures. The red channel contained the antibody-associated fluorescence detected by a fluorophore-conjugated secondary antibody.

For image processing, the green-channel image was first localized and normalized, followed by correction of background variation. Candidate cells were then generated, and their boundaries were refined or separated when necessary. The resulting cell masks defined the regions used for quantification of red-channel fluorescence, while adjacent pericellular regions served as background references. The green channel was used primarily for cell localization, whereas antibody-related signal evidence was derived mainly from the co-registered red channel.

Implementation details were as follows. Green-and red-channel intensities were independently rescaled to [0, 1] using the 1st and 99.7th percentiles, and images were downsampled only when the maximum dimension exceeded 1 024 pixels. For cell localization, the green channel was smoothed with a Gaussian filter (sigma=0.8), corrected by subtracting a broad Gaussian background (sigma=18), and segmented using layered hysteresis thresholding followed by distance-transform watershed. At a 768-pixel reference width, candidate cell areas were 35–3 600 pixels, the minimum seed distance was 8 pixels, and a 3-pixel border margin was excluded; these geometric values were scaled according to image resolution. The low threshold was max(0.36 × Otsu, 42nd percentile, 0.012), and the high threshold was max(0.82 × Otsu, 78th percentile, 0.06).

### Model development and evaluation

Model architecture was a three-class ExtraTreesClassifier. The 3 309-dimensional record-level input combined red-channel measurements within the mapped cell masks and local pericellular background rings, cell-distribution and top-versus-bottom contrasts, connected-component containment and spillover descriptors, mask morphology, segmentation quality, red-green spatial correspondence, and one-hot indicators for antibody and specimen type. Missing and infinite predictors were replaced with zero. The classifier used 325 trees, Gini criterion, square-root feature subsampling, unrestricted tree depth, minimum split and leaf sizes of 2 and 1, no bootstrap sampling, balanced class weighting, random_state=20260705, and 10 parallel jobs.

Training used only the 2 763-record development dataset. Five-fold stratified cross-validation used shuffling and random_state=20260705, with class-balanced sample weights in each fit. Out-of-fold probabilities were used to estimate the deterministic post-processing rules, after which the classifier was refit on all development records. Global and antibody-specific threshold features were estimated from the development records for positive-like versus negative and positive versus weak-positive/equivocal contrasts. The threshold-feature bank was estimated once from the complete development set rather than re-estimated within each cross-validation fold. The 499 reader-study records were excluded from model fitting and rule estimation but were used to compare candidate pipelines and for the reported evaluation.

The base label was the class with the largest of the three ExtraTrees probabilities. Development-derived rules were then applied to weak-positive/equivocal predictions. For AChR, post_achr_positive_score >0.6206 promoted a prediction to positive when P(negative) ≤0.75, whereas a score below 0.6206 demoted it to negative when P(positive) ≤0.25. For GAD65, post_gad65_strong_score >0.1596 promoted a prediction to positive when P(negative) ≤0.75. The rule search selected no active AQP4 or MOG spillover-demotion rule in this run. A gray-zone flag was calculated for non-negative predictions when the probability margin was ≤0.05 or normalized entropy was ≥0.55, but this flag did not replace the integrated three-class label used for the reported performance results.

No Platt scaling, isotonic regression, or other formal probability-calibration procedure was applied; model probabilities were used for class selection and rule guards and were not interpreted as calibrated clinical probabilities. The archived locked runtime specified NumPy 1.26.4, pandas 2.1.4, SciPy 1.16.1, scikit-learn 1.4.2, scikit-image 0.22.0, OpenCV 4.10.0.84, and matplotlib 3.8.0.

### Visual overlay generation

Detected cell boundaries were overlaid onto the corresponding red antibody-staining image, with the original fluorescence channels retained for visual review. The overlay provided a spatial reference for the segmented cellular regions. The complete image-processing and inference pathway is summarized in Supplementary Fig. S1.

### Reader study

Two readers were classified as experts and two as novices. Expert readers were defined as laboratory professionals with ≥3 years of experience in neurological autoantibody testing. Novice readers had <1 year of experience in neurological autoantibody CBA interpretation and had received brief standardized training before the reader study.

All four readers independently classified the 499 records as negative, weak positive/equivocal, or positive in two sequential sessions. In the first session, readers evaluated the original fluorescence images without access to the AI-generated category or overlay. After a washout interval of two weeks, the same records were presented in a newly randomized order for the second session, during which the corresponding AI overlay and model-generated category were available together with the original fluorescence images. Throughout both sessions, readers were blinded to clinical information, reference labels, the assessments of the other readers, and their own previous ratings. The study did not use a crossover design or a parallel control group.

Reader classifications were compared within each reader between sessions, with results also summarized separately for experts and novices. Reader-study outcomes were the proportion of ratings that changed between sessions, agreement between each reader and the model, agreement within the expert and novice pairs, complete four-reader agreement, overall pairwise agreement, the number of 2:2 ties, and Fleiss kappa. Changes were classified as moving directly to the model category, moving one ordinal step toward the model category, or moving away from the model category.

### Reference labels and model-performance measures

For standalone model evaluation, reference labels for the 499 records were derived from the four readers’ first-session votes without expertise-based weighting. The most-voted category was used; in 2:2 ties, archived labels corresponded to the lower category (negative < weak-positive/equivocal < positive). Tied records were included in model evaluation and retained as ties in reader-agreement analyses. This reader-derived reference was not an independent analytical or clinical gold standard. Clinical information and orthogonal laboratory results were unavailable for uniform adjudication of discordant records.

Model measures were three-class agreement with the project reference, balanced accuracy, binary agreement for negative versus non-negative classifications, and class-specific recall. Balanced accuracy was defined as the arithmetic mean of recall for the negative, weak-positive/equivocal, and positive classes.

## Statistical analysis

Analyses were descriptive at the record-reader level. Each experience group contributed 998 paired ratings (499 records × two readers), for 1 996 paired ratings overall. Wilson score 95% CIs were calculated for binomial model-performance proportions when numerators and denominators were available. Balanced accuracy, pairwise agreement, and Fleiss kappa are reported as descriptive point estimates without CIs. Reader change rates were not hypothesis-tested because ratings were clustered within records and readers and the study lacked randomization, a control condition, and an independent reference standard. Reporting was informed by DECIDE-AI, STARD-AI, and CLAIM 2024._12,13,14_

### Use of artificial intelligence

This manuscript was revised using generative AI tools for language editing and clarity. The authors take full responsibility for the scientific content, accuracy, and interpretation, in accordance with the journal policy.

## Results

### Study dataset

The development dataset comprised 2 763 records. Of 500 records identified for held-out evaluation, 499 were complete and included in the analysis. The evaluation set covered 14 antibody targets and contained 258 negative, 73 weak-positive/equivocal, and 168 positive reference labels (Fig. 1 and Table 1). Four readers each evaluated these 499 records in two sessions, providing 1 996 paired reader-record observations.

### Held-out model performance

Three-class agreement between the selected model and the reader-derived project reference was 85.17% (425/499; 95% confidence interval (CI), 81.78%–88.02%), and balanced accuracy was 8 77.27%. Binary agreement for negative versus non-negative classifications was 93.39% (466/499; 95% CI, 90.86%–95.25%). Recall was 93.02% (240/258; 95% CI, 89.24%–95.54%) for negative records, 50.68% (37/73; 95% CI, 39.47%–61.83%) for weak-positive/equivocal records, and 88.10% (148/168; 95% CI, 82.33%–92.16%) for positive records (Table 2).

**Table 2.** Standalone model agreement with the reader-derived reference in the 499-record held-out set. The reference was derived from first-session reader ratings and was not an independent analytical or clinical gold standard. Confidence intervals are Wilson score intervals.

| Metric | Estimate (95% CI) | Numerator/denominator or definition |
| --- | --- | --- |
| Three-class agreement with project reference | 85.17% (81.78%–88.02%) | 425/499 |
| Balanced accuracy | 77.27% | Mean recall across the three classes; CI not available |
| Negative vs non-negative agreement | 93.39% (90.86%–95.25%) | 466/499 |
| Negative recall | 93.02% (89.24%–95.54%) | 240/258 |
| Weak-positive/equivocal recall | 50.68% (39.47%–61.83%) | 37/73 |
| Positive recall | 88.10% (82.33%–92.16%) | 148/168 |

The weak-positive/equivocal class had the lowest recall. These estimates represent agreement with a reader-derived project reference and should not be interpreted as diagnostic sensitivity or specificity against disease status.

The complete 3 × 3 confusion matrix is shown in Fig. 2a. Most errors involved the weak-positive/equivocal class: 28/73 records were classified as positive and 8/73 as negative. Target-specific performance is reported descriptively in Fig. 2b because several antibody subgroups were small or lacked one or more reference classes. The most frequent antibody-specific error patterns were acetylcholine receptor (AChR) weak-positive/equivocal to positive (n=6), aquaporin 4 (AQP4) negative to positive (n=5), AQP4 weak-positive/equivocal to positive (n=5), contactin-1 (CNTN1) weak-positive/equivocal to positive (n=5), and N-methyl-D-aspartate receptor (NMDAR) positive to negative (n=5); these counts were not adjusted for subgroup size.

**Fig. 2.**
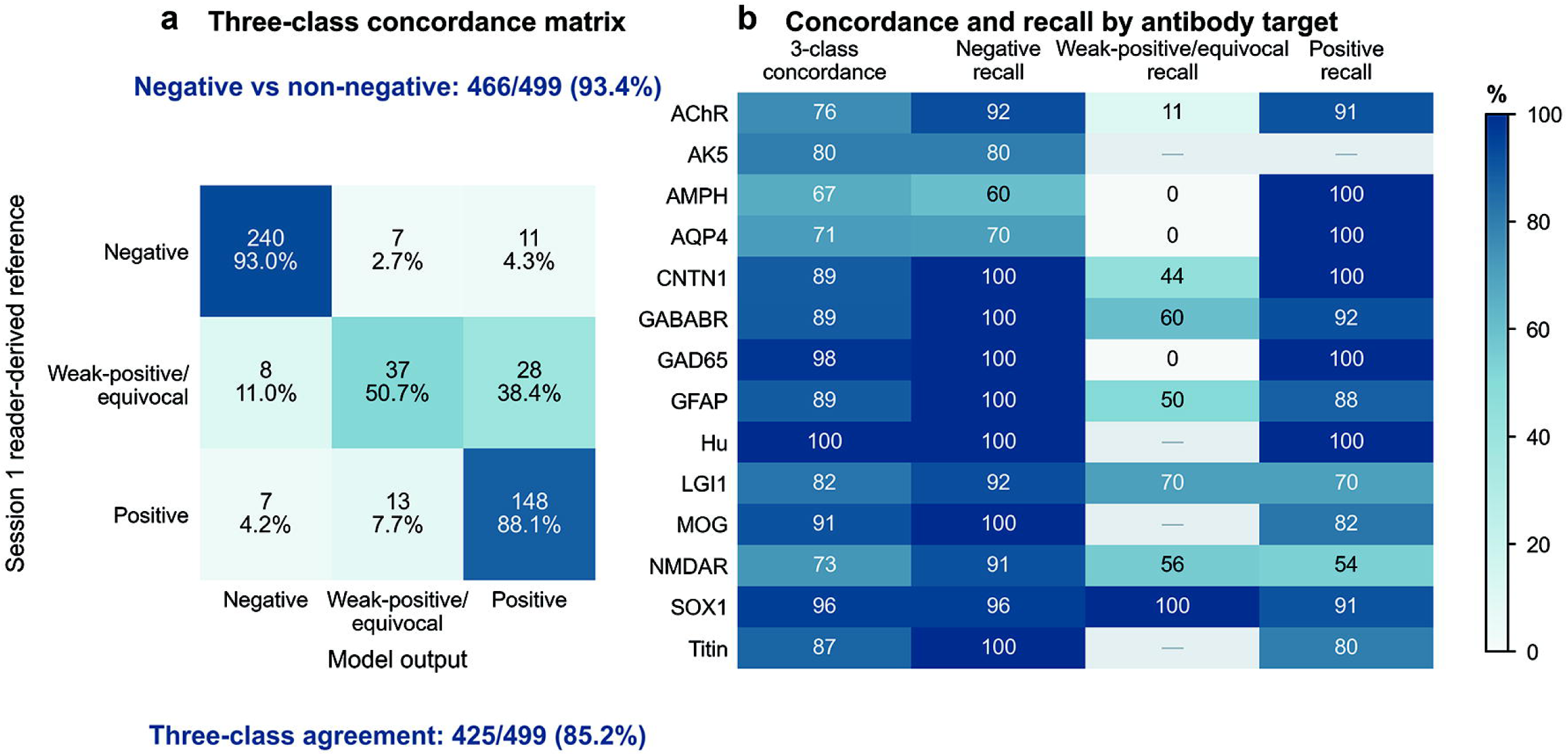
Standalone model concordance with the reader-derived reference. a, Three-class confusion matrix for the 499-record held-out model-selection set. Rows indicate the reader-derived reference and columns indicate model output; percentages are normalized within reference class. b, Descriptive three-class accuracy and class-specific recall by antibody target. Dashes indicate that the corresponding reference class was absent. Subgroup results are descriptive because several groups were small and uneven.

### Visual overlays

The workflow identified cell boundaries in the green channel and mapped them onto the red antibody-staining channel. Representative negative, weak-positive/equivocal, and positive records are shown in Fig. 3. The overlays show the segmented cellular regions while retaining the original fluorescence pattern.

**Fig. 3.**
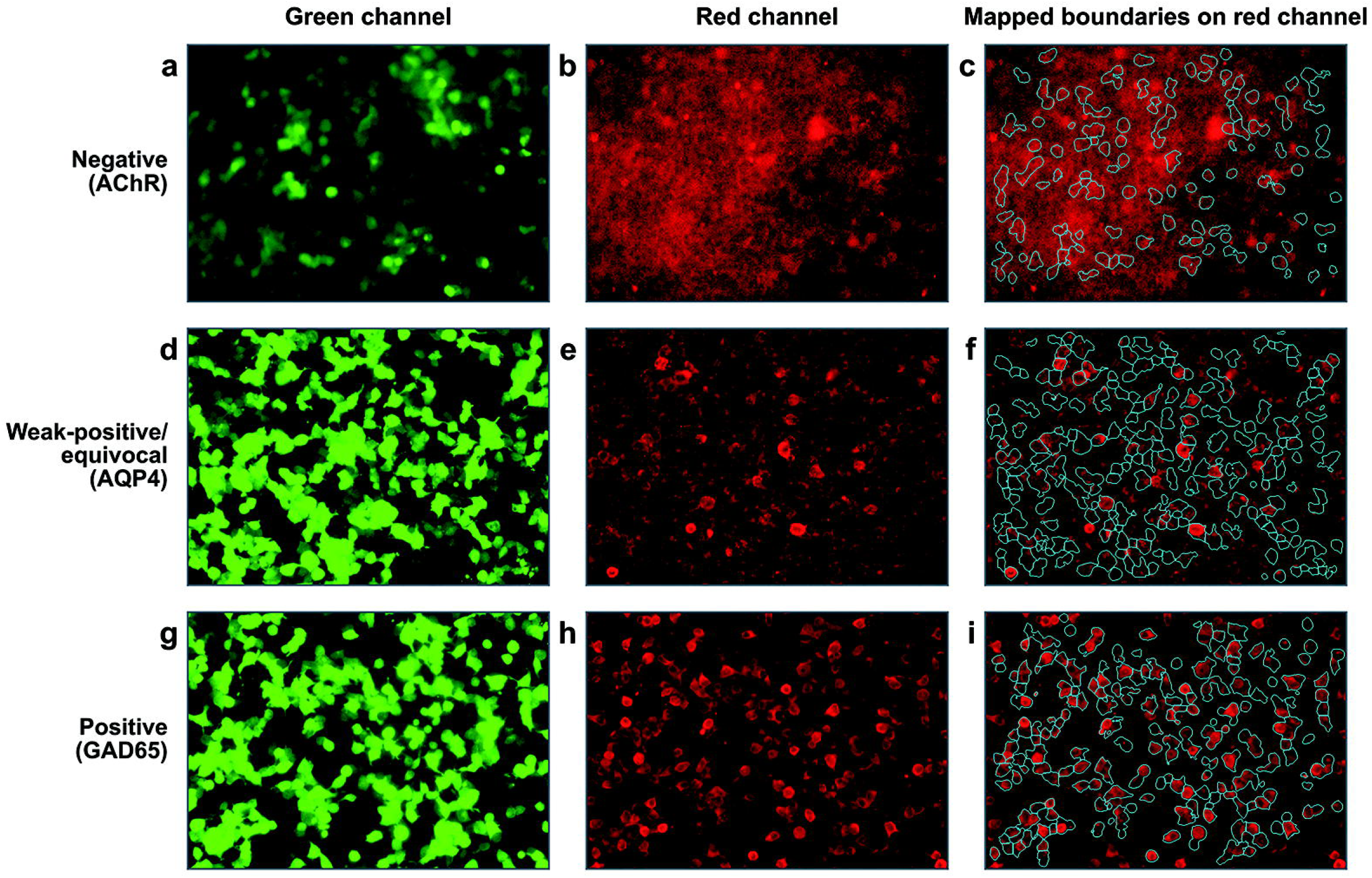
Representative paired-channel cell-based assay (CBA) images and mapped cell boundaries. a–c, Representative negative acetylcholine receptor record. d–f, Representative weak-positive/equivocal aquaporin-4 record. g–i, Representative positive glutamic acid decarboxylase 65 record. Within each row, the green structural channel is followed by the corresponding red antibody-staining channel and the mapped cell-boundary overlay.

### Global feature importance

Global descriptive mean decrease in impurity (MDI) analysis showed that feature importance was distributed across several related feature families (Supplementary Fig. S2). Antibody-specific threshold features contributed 32.6% of total MDI, red-channel evidence features 27.0%, and fast-cell threshold features 21.7%. The highest-ranked individual feature accounted for 1.54%, while the top 500 ranked features accounted for 55.4% of cumulative MDI. These values describe how the fitted ensemble used the available predictors and do not establish causal or biological importance.

### Overall changes in reader interpretation

Across 1 996 paired ratings, 73 (3.7%) changed in the second, AI-assisted session. The four-reader majority category changed for 27 records (5.4%), most commonly from weak positive/equivocal to positive (20 records). The distribution of changed ratings by reader experience is shown in Fig. 4c.

**Fig. 4.**
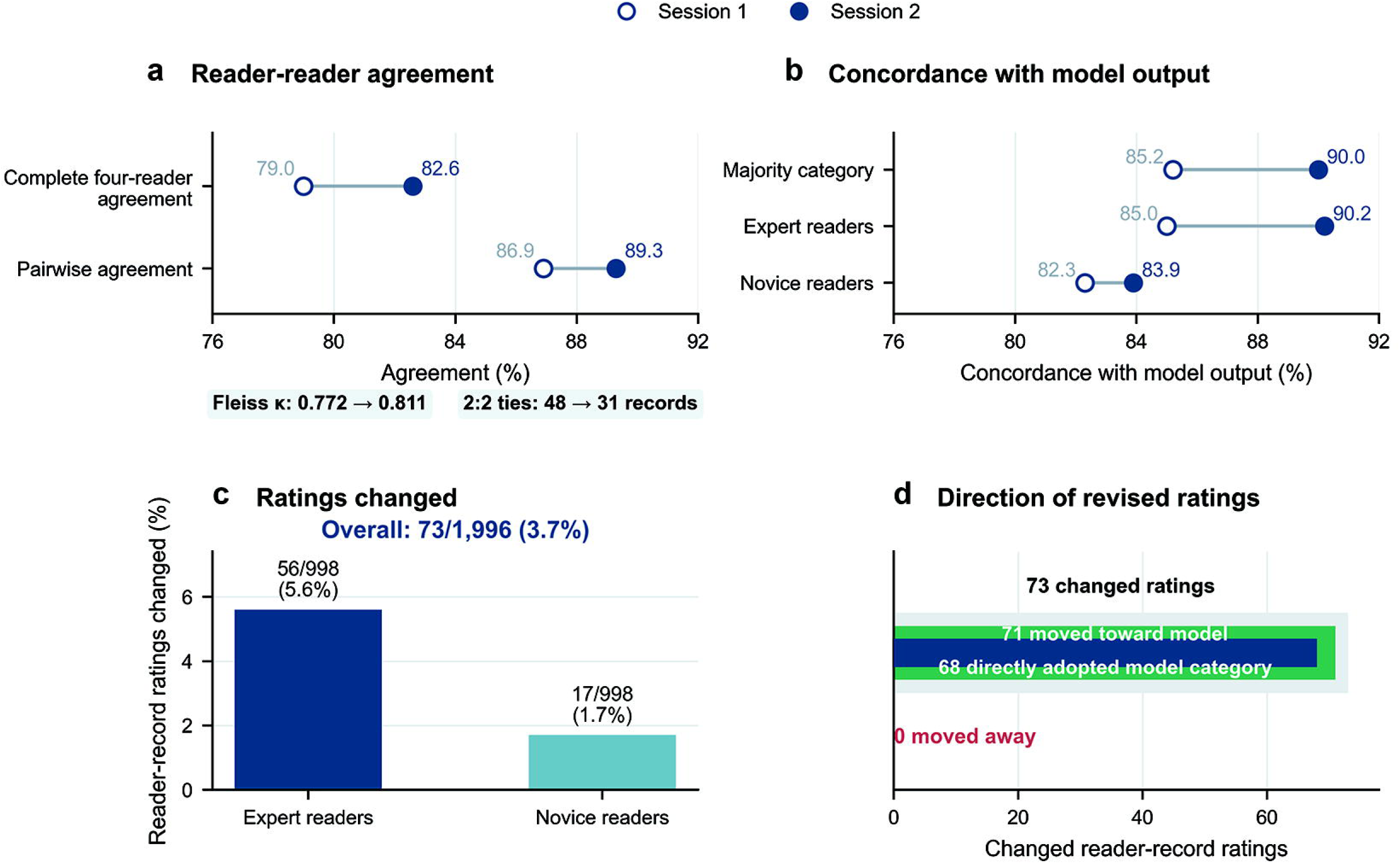
Reader-study outcomes in unassisted and AI-assisted sessions. a, Complete four-reader and pairwise agreement in sessions 1 and 2. b, Concordance between reader categories and model output. c, Proportion of ratings changed by expert and novice readers. d, Direction of the 73 revised ratings relative to the model category. The study included 499 records, four readers, and 1 996 paired reader-record observations; all comparisons are descriptive.

Complete four-reader agreement increased from 79.0% to 82.6%, pairwise agreement from 86.9% to 89.3%, and Fleiss kappa from 0.772 to 0.811; 2:2 ties decreased from 48 to 31. Majority-category agreement with the model increased from 85.2% to 90.0%. Of the 27 majority-category changes, 24 moved directly to the model category and none moved away (Fig. 4a,b and Supplementary Table S1).

### Reader-study outcomes by experience level

Experts changed 56/998 ratings (5.6%), compared with 17/998 (1.7%) among novices. Expert-model agreement increased from 85.0% to 90.2% and expert-pair agreement from 95.0% to 96.2%.

Novice-model agreement increased from 82.3% to 83.9%, while novice-pair agreement decreased from 90.6% to 90.0%. The most frequent expert change was weak positive/equivocal to positive (43 ratings); for novices it was positive to weak positive/equivocal (eight ratings). These experience-stratified patterns are summarized in Fig. 4b,c.

### Direction of rating changes

Of the 73 changed ratings, 68 (93.2%) moved directly to the model category. Including these 68 ratings, 71 (97.3%) moved closer to the model category; none moved farther away. These changes describe alignment with the model, and their clinical correctness was not established by an independent reference standard (Fig. 4d and Supplementary Table S2).

## Discussion

Changes in repeated image classification during the assisted session differed between the two experience groups. Experts revised more ratings, particularly at the weak-positive-to-positive boundary, whereas novices changed fewer ratings and did not show higher within-pair agreement. Interpretive expertise influences neural antibody immunofluorescence assessment.^5,11^

AI-assisted interpretation has been studied in skin cancer recognition, liver histopathology, and prostate biopsy grading^15,16,17^. In our CBA setting, the model had its lowest recall in the weak-positive/equivocal class, where additional review may be most valuable. A cell-boundary overlay can show the spatial regions used for image quantification, but it does not resolve ambiguity in low-intensity, focal, or background-confounded fluorescence. Clinical information and orthogonal laboratory findings were not uniformly available for adjudication. The study therefore cannot determine whether reader disagreement reflected differing interpretive thresholds, uncertainty, or other factors.

The overlay provided a visual reference by showing the boundaries of GFP-marked transfected cells on the antibody-staining channel. Overall reader agreement increased in the assisted session, while novice within-pair agreement decreased. These findings describe consistency of image review. They do not establish improvement in diagnostic accuracy or demonstrate that the boundary display explains the contribution of individual features to a model prediction.

Almost every revised rating moved toward the model, and none moved away. This pattern is compatible with model anchoring, although the present design cannot determine whether individual changes were correct. Experimental studies show that inaccurate AI advice can reduce clinician performance and that explanations do not consistently protect users from biased model recommendations^18,19^. Higher reader-model agreement therefore should not be equated with higher diagnostic accuracy.

The greater expert change rate does not establish greater benefit, because the study lacked an independent outcome standard. Whether experts and novices use the AI-assisted display differently requires evaluation in a larger reader sample. Training approaches that combine AI-assisted displays with grading criteria and feedback also require prospective evaluation.

A cautious workflow would present the overlay and model category only after an independent assessment of the original channels. Weak-positive, focal-positive, high-background, atypical, or model-reader discordant cases should remain subject to expert review using the original images and, when available, orthogonal laboratory results and clinical context.

Such staged use may preserve attentional guidance while limiting premature anchoring. Prospective evaluation should measure override behavior and the effect of model errors, consistent with DECIDE-AI principles^12^.

This study has several limitations. First, the reader-derived reference created incorporation and circularity concerns. Second, despite a two-week washout interval and randomized image order in the second session, the fixed sequence of unassisted followed by assisted reading, without a crossover design or parallel control group, means that effects of the combined AI display cannot be separated from repeat-reading effects or attributed to either the overlay or model category alone. Third, only two experts and two novices participated, limiting generalizability across readers with different levels of experience. Fourth, exact duplicate images were excluded, but patient-level separation between development and held-out datasets was not established. Fifth, because the held-out set was used to compare candidate pipelines, it was not a strictly untouched external test set. Sixth, no separate probability-calibration procedure was used, and some acquisition parameters (magnification, exposure, and pixel dimensions) were unavailable. Finally, the added confusion matrix and antibody-target analyses remain descriptive; prespecified specimen-and laboratory-level analyses were unavailable, and several target groups were small. These limitations restrict reproducibility, causal inference, and generalizability.

The AI-assisted session was associated with higher reader agreement and model-directed rating changes, but whether these changes improved interpretation remains unknown, particularly for weak-positive/equivocal records. Future studies should use patient-level data separation, independent external test sets, and reference standards incorporating orthogonal testing and clinical information. Randomized crossover or parallel reader studies with larger samples should measure accuracy and responses to incorrect AI advice to distinguish useful guidance from model anchoring and repeat-reading effects. Complete acquisition documentation, calibration assessment, and prespecified subgroup analyses would further strengthen evaluation, in line with DECIDE-AI, STARD-AI, and CLAIM 2024.^12,13,14^

## Data availability

The de-identified data supporting this study are available from the corresponding authors upon reasonable request, subject to applicable privacy and ethics requirements.

## Code availability

The code used in this study is available from the corresponding authors upon reasonable request.

## Author contributions

Zhihui Qi and Zijian Cao: Conceptualization, Methodology, Investigation, Data curation, and Formal analysis. Hongyi Tan and Boxiang Tang: Software, Image processing, Model development, and Technical validation. Samuel X Shi, Ruoyong Jia, Yang Yao, Jingshan Chen and Zongjiu Zhang: Clinical sample collection, CBA testing, Image acquisition, Data curation, and Clinical interpretation. Jiahao Wan: Conceptualization, Methodology, Software, Formal analysis, Visualization, and Writing-original draft. Pei Zheng: Conceptualization, Supervision, Project administration, Funding acquisition. All authors reviewed and approved the final manuscript.

## Ethics approval and consent to participate

The specimens had originally been submitted for routine patient care and were jointly provided by the neuroimmunology laboratories of Tianjin Medical University General Hospital and Beijing Tiantan Hospital, Capital Medical University. No specimens were collected specifically for this study. The study was approved by the Ethics Committee of Tianjin Medical University General Hospital (IRB No. IRB2023-KY-060) and the Science and Technology Ethics Committee (Institutional Review Board) of Tsinghua University (Project No. THIU-01-2025-1020). Given the retrospective nature of the study and the use of previously collected, de-identified specimens and data, the requirement for written informed consent was waived by the ethics committees. The study was conducted in accordance with the Declaration of Helsinki.

## Supplementary information

The accompanying files contain Supplementary Figures S1 and S2 and Supplementary Tables S1 and S2.

## Data Availability

All data produced in the present study are available upon reasonable request to the authors. Due to patient-privacy constraints, raw clinical sample-derived datasets cannot be publicly deposited.

