## Supplementary_Information for "AI-Assisted Interpretation of Neurologic Autoantibody Cell-Based Assays"

Supplementary Fig. S1. Technical pipeline. a, cell-based assay (CBA) images were acquired as paired structural and antibody channels. Cells were localized from the structural channel, and the corresponding boundaries were transferred to the antibody channel. A total of 3 309 record-level features were supplied to an ExtraTrees classifier with 325 trees and deterministic decision rules, producing locked three-class inference. b, During the second reader session, the boundary overlay and model-generated category were presented with the original images.

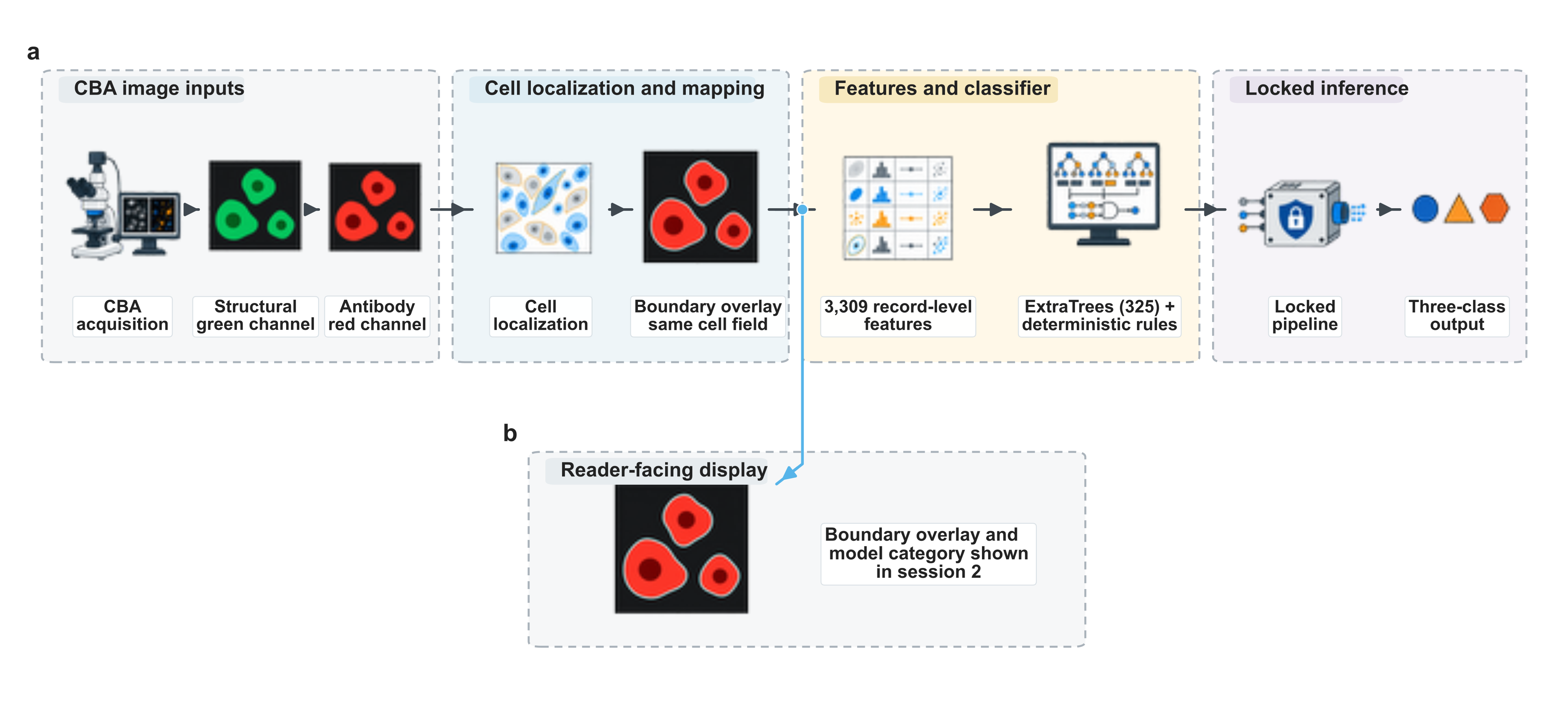

Supplementary Fig. S2. Global descriptive feature importance in the fitted ExtraTrees model. a, Share of total mean decrease in impurity (MDI) importance by feature family. b, The 12 highest-ranked individual features. c, Cumulative MDI importance across ranked features. MDI values describe feature use within the fitted ensemble and should not be interpreted as causal or biological effects.

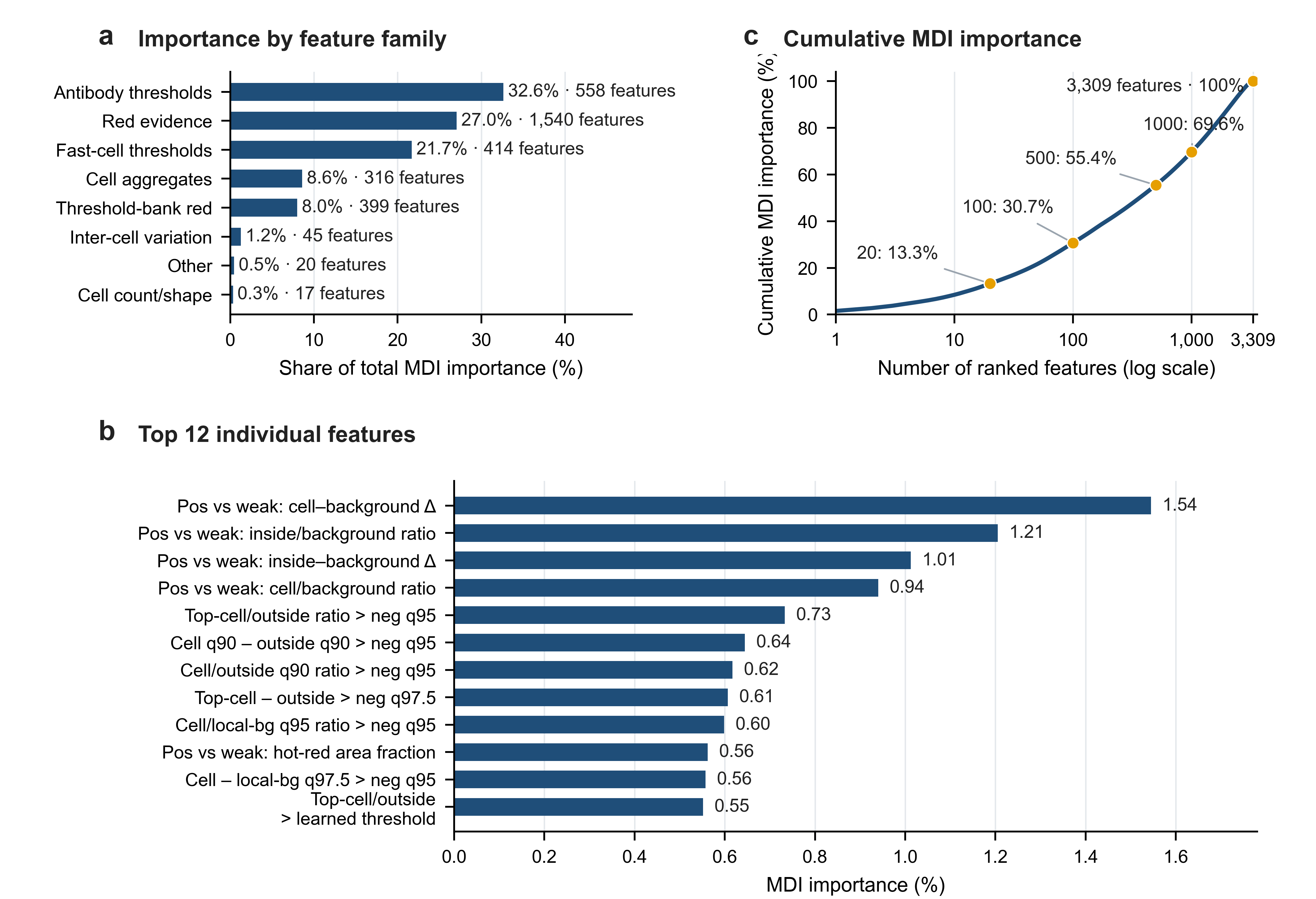

**Supplementary Table S1. Overall reader agreement in unassisted and AI-assisted sessions. Values are descriptive for the 499-record paired reader study. Pairwise agreement and Fleiss kappa are reported without confidence intervals.**

| **Metric** | **Session 1** | **Session 2** | **Absolute change** |
| --- | --- | --- | --- |
| Complete four-reader agreement | 79.0% (394/499) | 82.6% (412/499) | +3.6 percentage points |
| Pairwise agreement | 86.9% | 89.3% | +2.4 percentage points |
| Fleiss kappa | 0.772 | 0.811 | +0.039 |
| Tied records | 48 | 31 | −17 |
| Majority-category agreement with model | 85.2% | 90.0% | +4.8 percentage points |

**Supplementary Table S2. Grading changes and experience-stratified outcomes. Expert and novice groups each included two readers. Arrows indicate change from session 1 (original images only) to session 2 (original images, boundary overlay, and model category). 'Not stratified' indicates that the archived analysis did not provide the corresponding experience-specific count.**

| **Metric** | **Overall** | **Expert readers** | **Novice readers** |
| --- | --- | --- | --- |
| Changed ratings | 73/1 996 (3.7%) | 56/998 (5.6%) | 17/998 (1.7%) |
| Agreement with model | Not applicable | 85.0% → 90.2% | 82.3% → 83.9% |
| Within-group agreement | Not applicable | 95.0% → 96.2% | 90.6% → 90.0% |
| Most frequent change | Weak positive/equivocal → positive (46) | Weak positive/equivocal → positive (43) | Positive → weak positive/equivocal (8) |
| Changed directly to model category | 68/73 (93.2%) | Not stratified | Not stratified |
| Moved toward model category | 71/73 (97.3%) | Not stratified | Not stratified |
| Moved away from model category | 0 | Not stratified | Not stratified |
